# Improving palliative care for babies, children, young people and adults from ethnically diverse communities: a qualitative multiple case study

**DOI:** 10.64898/2026.07.01.26356998

**Authors:** Lesley Dunleavy, Sam Gould, Gemma Clarke, Natalie Cotterell, Sabrina Bajwah, Catherine Evans, Lorna Fraser, Sarah Mitchell, Nancy Preston, Catherine Walshe

## Abstract

**Background:** Palliative care services appear less able to reach people from ethnically diverse communities and if they do, people from these communities’ report having different and often poorer experiences. The barriers to access are well described so research investigating potential solutions is needed.

**Aims:** To understand how improved palliative care outcomes for people from ethnically diverse communities have or could be achieved, and what contextual issues have influenced these outcomes.

**Methods:** Qualitative multiple case study. The cases were defined as areas across England with services providing generalist or specialist palliative care to adults and/or children. Interviews were conducted with patients, family carers, parents (from ethnically diverse communities), health and social care professionals. Data were analysed using thematic framework analysis.

**Findings:** Cases (n=6) included 71 participants. Five solution focused themes were identified; how the conditions for culturally and spiritually safe care are created; engagement and trust building between ethnically diverse communities and the providers that serve them as a mechanism to promote access; workforce composition and diversity helping to bridge the gap between ethnically diverse communities and services; how communication practices enable equitable care for people who have English as an additional language and organisational commitment and partnership as drivers of sustainable change.

**Conclusions:** Equitable access to quality palliative care is not a marginal policy issue with the economic and moral argument for change strong. Care systems need to recognise, partner with, and build upon existing community strengths. Change requires intent, accountability, leadership and the reallocation of attention and responsibility.

**Highlights:**

- Environmental changes and flexible care enable people to feel safe and respected.
- Using an asset-based community engagement approach is useful to improve access.
- Local staff can bridge the gap between services and ethnically diverse communities.
- Access to interpreters and translated material is imperative for equitable care.
- Sustainable change requires organisational commitment and partnership working.

## Introduction and background

The principle that everyone should have equitable access to high-quality palliative care is firmly embedded in health policy (NHS England, 2022a). Yet despite increased awareness, service development, and cultural competency initiatives, persistent inequalities remain for people from ethnically diverse communities (Aker et al., 2024; LoPresti et al., 2016; Tobin et al., 2021). Palliative care services are less likely to reach individuals from these communities, and when access does occur, experiences are often different and comparatively poorer (Chukwusa et al., 2025; National Audit of Care at the End of Life (NACEL), 2025); Selman et al. (2023).

The demographic context in which these disparities occur is changing rapidly. Populations are becoming increasingly diverse, with individuals from ethnically diverse communities being born, growing, ageing, and living with complex, long-term, and life-limiting conditions (Catney, 2016; Hayanga et al., 2023). Consequently, demand for palliative care among babies, children, young people, and adults from ethnically diverse communities is rising, with a higher prevalence of life-limiting conditions in some populations (Aker et al., 2024; Fraser et al., 2021; Odd et al., 2024). However, utilisation of palliative care services among ethnically diverse communities often remains below levels expected based on need (Tobin et al., 2021). This mismatch raises critical questions about whether current healthcare systems are designed to meet the needs of increasingly multicultural societies.

These inequities have major implications. Palliative care is fundamentally concerned with dignity, comfort, and support during periods of profound vulnerability. Where individuals lack awareness of specialist palliative care services or face barriers to accessing generalist or specialist provision, opportunities to alleviate pain, support families, and improve wellbeing may be missed, with adverse consequences for quality of life and death (Ip et al., 2026; Richards, 2022). Inequitable access also amplifies existing health disparities, reduces support for families during crises, and is associated with poorer bereavement outcomes. From a system perspective, inequitable patterns of service use can distort planning and commissioning, perpetuating models of care that reflect the needs and expectations of a limited segment of the population (Mitchell et al., 2024; Rhodes et al., 2022).

The drivers of these inequities are complex and multifactorial. Previous research has highlighted the influence of cultural and religious expectations, differing understandings of palliative care, and variations in family roles and decision-making practices across communities (Shabnam et al., 2022). Language and communication barriers further shape access and experiences (Dookie & Martin, 2025; Dressler et al., 2021). In addition, structural and systemic factors play a critical role, including inconsistent referral patterns, historical and ongoing mistrust of healthcare institutions, and service designs that reflect majority norms (Holder et al., 2024). Importantly, relatively few initiatives explicitly address structural racism, which may be embedded within both community and organisational contexts, including within palliative and end-of-life care systems (J. Hussain et al., 2021).

In summary, palliative care services frequently provide inadequate (or missing) care for people from ethnically diverse communities, within a context characterised by complexity and intersecting barriers. While the existence of inequities is well documented, there remains limited articulation of policy-relevant solutions. The aim of this study is to explore, in context, potential solutions to the challenges and barriers shaping how people from ethnically diverse communities access, use, and are offered palliative care.

When undertaking this study we sought to avoid deficit-oriented terminology that positions groups primarily in relation to a dominant majority. We have chosen to use the term ethnically diverse communities as it reflects diversity as a strength and acknowledges the cultural knowledge, networks, resilience, and contributions present within communities.

## Methods

### Aim

To understand how improved palliative care outcomes for people from ethnically diverse communities have or could be achieved, and what contextual issues have influenced these outcomes.

### Design

A mixed-method multiple case study research strategy (Yin, 2018) with a critical realist lens applied throughout the study (Bhaskar, 2013). Theoretical propositions guided the study design, data collection and analysis process reflecting a critical realist stance (Yin, 2018). Initial theoretical propositions were developed from a review that preceded this study and are presented in supplementary materials (Supplementary Materials 1).

### Case definition

The boundary of the case was defined as a specified healthcare geography (place) in England within which there were either current or recent initiative/s of any form aimed at improving access to, or experience of, palliative care to people from ethnically diverse communities. Our definition of initiative was broad, including any activities provided to adults and/or children/young people. Palliative care refers to services provided by specialist or generalist palliative care professionals.

### Selection of cases

Case sampling was informed by our initial theoretical propositions and took account of intersectionality, interventions or initiatives and populations with proportionately large ethnically diverse communities. One case explicitly focused on paediatric populations/care/initiatives, although paediatric services could form part of any of the cases. Contextually only 74.4% of residents in England and Wales identify as white British with the rest identifying as Asian/Asian British (9.3 %), White Other (6.2%), Black/Black British/Caribbean/African (4.0 %), Mixed or Multiple Ethnic Groups (2.9%) and Other Ethnic Group (2.1%) (Office for National Statistics (ONS), 2022).

### Participants within the cases

People with a life limiting illness/conditions and/or palliative care needs from ethnically diverse communities (including children 12-15 with capacity) residing within, or receiving care within, the case geography and their family/informal carers/parents (including bereaved carers/parents) were eligible. Participants also included relevant stakeholders from or known to the initiatives selected as cases for this study including health and social care professionals, service managers and commissioners. They could work within primary care, acute care, hospices, community, and social care or in the third sector or related communities.

### Sampling and recruitment within the cases

Potential patient and family carer participants were purposively sampled within cases to maximise variety based on different characteristics such as gender, diagnosis, ethnicity and age. Potential stakeholder participants were selected purposively by the research team, following discussion with key case contacts, identified as people who could give rich data on the phenomena under study.

A preliminary assessment of what documents or publicly available routine data were available online was made for each ‘case’. Study participants were asked to identify and share documents that were relevant to the study aims as appropriate.

Participants were recruited via care organisations within the case who provided information to potential participants that met eligibility criteria. Additional recruitment methods included circulating advertisements via posters and social media.

### Data collection

Data collection took place sequentially over 16 months (October 2024-January 2026). Data collection included in-depth interviews and the collection of relevant documentary evidence. Interviews took place either online (using Microsoft Teams), via the telephone, or in-person and were digitally recorded, transcribed and anonymised. The interviews were carried out by (researcher initials) experienced palliative care researchers who identified as White British with (researcher initials) also having nursing experience. Translated materials and/or professional interpreters were available. The topic guide was not a fixed schedule but typically included the type of support received, their views and experiences of these services (including instances of culturally/religiously congruent care), accessibility issues, wider support networks, their views of the type of services needed to meet the needs of ethnically diverse communities. For stakeholders, it typically included the description of initiatives or activities that are in place within their service/organisation and wider local area to meet the needs of ethnically diverse communities living with a life limiting illness, any implementation issues including any barriers and facilitators and recommendations for shared learning.

### Data analysis

A Framework Analysis approach facilitated analysis between cases using a matrix approach and within and cross case pattern matching (Dunleavy et al., 2023; Walshe et al., 2008). The approach involves a systematic process of sifting, charting, and sorting material following five key stages: familiarisation, identifying a thematic framework, indexing, charting, and mapping and interpretation (Ritchie & Lewis, 2003). The initial thematic framework was formed from our initial theoretical propositions and subsequently iterated and developed.

Cross case pattern matching followed to identify thematic factors associated with improved palliative and end of life care outcomes. Data analysis was carried out by (LD, SG, GC, NC, CW) as described above with input and guidance from the wider team (SB, CE, LF, SM, NP). The wider team are experienced palliative care academic researchers and clinicians with the majority identifying as White British. A Patient and Public Involvement and Engagement group (PPIE) was set up specifically to guide this project which included reviewing study findings.

### Ethical issues

This study was reviewed and approved by the Health Research Authority and North West-Liverpool Central Research Ethics Committee (REC reference number 24/NW/0214, IRAS ID 344205, 22^nd^ July 2024. Assent to contact potential participants was obtained prior to contact. Informed consent was gained prior to the interviews.

## Findings

Descriptions of the six cases and their participants included in the study are summarised in table 1 (see supplementary file 1 for additional participant demographics). The findings are presented as a cross-case analysis and are presented as five overarching themes:

- How the conditions for culturally and spiritually safe care are created.
- Engagement and trust building between ethnically diverse communities and the providers that serve them as a mechanism to promote access.
- Workforce composition and diversity helping to bridge the gap between ethnically diverse communities and services.
- How communication practices enable equitable care for people who have English as an additional language.
- Organisational commitment and partnership as drivers of sustainable change.

**Table 1.** Description of the cases and participants.

|  | **Case One** | **Case Two** | **Case Three** | **Case Four** | **Case Five** | **Case Six** |
| --- | --- | --- | --- | --- | --- | --- |
| Geography/<br>Demography of the<br>case*<br>* All data are<br>rounded. Data given<br>on ethnically diverse<br>communities where<br>% greater than 5%<br>except for<br>mixed/multiple. | Large Town<br>Population<br>**134,000**<br>**Ethnicity:**<br>36% Asian/Asian<br>British<br>61% White | Inner city<br>Population<br>**239,000**<br>**Ethnicity:**<br>22% Black/Black<br>British/Caribbean<br>11% Asian/Asian<br>British.<br>9% Other<br>54% White | Large City<br>Population<br>**526,000**<br>**Ethnicity:**<br>33% Asian/Asian<br>British<br>62% White | Small City<br>Population<br>**127,000**<br>**Ethnicity:**<br>21% Asian/Asian<br>British<br>73% White | Large City<br>Population<br>**348,000**<br>**Ethnicity:**<br>44% Asian/Asian<br>British<br>8% Black/Black<br>British/Caribbean<br>41% White | Inner city<br>Population<br>**319,000**<br>**Ethnicity:**<br>33% Asian/Asian<br>British<br>18% Black/Black<br>British/Caribbean<br>10% Other<br>35% White |
| Provider<br>organisations<br>participating | Hospital and<br>Children's Hospice | Hospital and Adult<br>Hospice | Hospital, Adult<br>Hospice and<br>Community Trust | Hospital, Adult<br>Hospice and<br>Children's Hospice | Adult Hospice,<br>Children's Hospice<br>and Community<br>Trust | Children's Hospice |
| Documents<br>collected | 3 (Newsletter,<br>Presentation,<br>Flowchart) | 1 (Published paper) | 0 | 1 (Service<br>Evaluation report) | 2 (Published paper<br>and study report) | 0 |
| **Health and Social Care Professional (including volunteers) participants** |  |  |  |  |  |  |
| Gender | F=7 | F=6<br>M= 2 | F=9<br>M= 2 | F=9<br>M= 1 | F=8 | F=3<br>M= 1 |
| Country of birth | UK=7 | UK=3<br>Non-UK=5 | UK=9<br>Non-UK=2 | UK=10<br>Non-UK=0 | UK=6<br>Non-UK=2 | UK=2<br>Non-UK=2 |
| Ethnic Group | White British= 4<br>Asian/Asian British<br>=3 | White British= 4<br>White other=2<br>Black, Black British,<br>Caribbean or<br>African=1<br>Other ethnic<br>group=1 | White British= 5<br>Asian/Asian British<br>=5<br>Other ethnic<br>group=1 | White British= 6<br>Asian/Asian British<br>=3<br>Black, Black British,<br>Caribbean or<br>African=1 | White British= 5<br>Asian/Asian British<br>=2<br>Black, Black British,<br>Caribbean or<br>African=1 | White British=1<br>Asian/Asian British<br>=2<br>Mixed/multiple<br>ethnic group= 1 |
| Main Language | English=7 | English=8 | English=9<br>Arabic=1<br>English & Urdu=1 | English=8<br>Gujarati=1<br>Gujarati and English=1 | English=5<br>Gujarati and English=2<br>Shona=1 | English=2<br>English/Cantonese=1<br>French=1 |
| Primary professional Role | Doctor=1<br>Nurse=3<br>Counsellor= 1<br>Family support worker=1<br>Volunteer=1 | Doctor=1<br>Nurse=3<br>Occupational therapist = 1<br>Psychologist=1<br>Spiritual care worker=1<br>Volunteer=1 | Doctor=3<br>Nurse=4<br>Spiritual care worker=1<br>Manager/admin=2<br>Link worker=1 | Doctor=1<br>Nurse=2<br>Family support worker=1<br>Spiritual care worker=1<br>Manager/admin=2<br>Educator=1<br>Volunteer=2 | Nurse=3<br>Family support worker=1<br>Volunteer=1<br>Manager/admin=1<br>Link worker=1<br>Key worker=1 | Nurse=1<br>Counsellor=1<br>Therapist=2 |
| Primary work Environment | Hospital= 4<br>Hospice= 3 | Hospital= 5<br>Hospice= 2<br>Community=1 | Hospital= 3<br>Hospice= 5<br>Community=3 | Hospital=3<br>Hospice= 6<br>Community=1 | Hospice= 6<br>Community=2 | Hospice=4 |
| Patient and family carer participants |  |  |  |  |  |  |
| Gender | M=1<br>F=1 |  | M= 4<br>F=5 |  | M= 4<br>F=6 | M= 1<br>F=1 |
| Age Group | 31-40=1<br>41-50=1 |  | 31-40=3<br>41-50=4<br>52-60=1<br>71-80=1 |  | 31-40=2<br>51-60=4<br>61=70=4 | 18-30= 1<br>41-50=1 |
| Country of birth | UK= 1<br>Non-UK =1 |  | UK=5<br>Non-UK=4 |  | UK=3<br>Non-UK=7 | Non-UK=2 |
| Ethnic Group | Asian/Asian British =2 |  | Asian/Asian British=6<br>Other ethnic group=1<br>White other=1<br>White British=1 |  | Asian/Asian British=8<br>Mixed/multiple ethnic group= 1<br>Black, Black British, Caribbean or African=1 | Black, Black British, Caribbean or African=1<br>White other=1 |
| Religion | Muslim=2 |  | Muslim=5<br>Hindu=3<br>Christian=1 |  | Hindu=6<br>Muslim=2<br>Christian=2 | Jewish=1<br>Christian=1 |
| Main Language | English=1<br>Urdu=1 |  | English=7<br>Arabic=1<br>Portuguese=1 |  | English=5<br>Hindi/English=1<br>Mandarin=1<br>Gujarati=1<br>Hindi=1<br>Urdu=1 | Ga=1<br>Romanian=1 |
| Total number of participants in each case | 9 | 8 | 20 | 10 | 18 | 6 |

### How the conditions for culturally and spiritually safe care are created

Creating an environment for culturally and spiritually safe care was imperative to facilitating equitable care. Implementing visible and practical changes to meet cultural and faith related needs and demonstrating flexibility in care provision enabled people to feel welcomed, safe and respected. Offering cultural and faith specific resources to patients and families and supporting religious rituals allowed staff to demonstrate that they cared about the persons culture and religion, recognised its importance and actively wanted to improve the care they delivered. It signalled inclusivity and support for patients and families in ways that were meaningful to them:

> *‘One of the nurses brought the lights. Sorry, the stars. And I thought that was wonderful. You know, they knew, (son’s name). This was potentially his last, you know, religious Eid, really. And they did everything on that level. So I was very pleased with that, you know,’* **Case five,** Parent (Bereaved), Children’s Hospice, Asian/Asian British

The availability of cultural and faith specific resources and policies within an organisation offered guidance for staff and demonstrated that a person’s needs have been actively considered:

> *‘What we provide for example is Prayer boxes for Muslim families, So we have their little like speakers and they basically repeat Quranic verses. And it’s it’s a detail, but it’s actually helping families.’* **Case two**, Doctor, Hospital, White (Other)

Flexible visiting arrangements and waiting spaces that could accommodate larger numbers of visitors were seen as important. They reflected the realities of diverse family practices and contributed to an environment where all families felt comfortable and accepted:

> *‘So when I saw how our IPU (inpatient unit) ward admin was with, you know, a large Asian family and I saw how normal she was’* **Case three,** Administrator, Adult Hospice, Asian/Asian British

Providing and offering inclusive resources and policies also demonstrated to staff and volunteers that the organisation was accepting, supportive and cared about their own cultural and spiritual needs.

Hospices worked to balance acknowledging their often-Christian heritage with creating environments in which people of all faiths felt welcome. Some participants suggested that hospice organisations could go further to create a more culturally safe environment by shifting away from Christian terminology and iconography:

> *‘One of the barriers that I do come across and it’s not, to be fair purely for minority ethnic backgrounds. But I think it probably has a disproportionate effect among minority ethnic background patients is that a lot of hospices have got quite Christian type names’* **Case one**, Doctor, Hospital, White British

### Engagement and trust building between diverse ethnic communities and the providers that serve them as a mechanism to promote access

Proactive community engagement between ethnically diverse communities and palliative care services was seen as a key means of improving access to palliative care. Community activities could provide valuable opportunities for mutual learning, support shared understanding, surface unmet communication needs such as concerns palliative care could intentionally hasten death, reduce fear and build trust between communities and palliative care services:

> *‘The primary goal is to listen to communities, provide education, and dispel the myths and taboos surrounding end of life care. We address the ethical, cultural and faith related issues around death and dying from a Muslim perspective.’* **Case one,** newsletter

Adopting and embracing an asset-based community engagement and development approach rather than looking for and filling gaps in service provision was seen as a useful strategy to improve access:

> *‘Don’t look for the gaps. You look for the strengths. So you’re treasure hunting. You’re going to find out where there is good stuff, and then you’re building on top of that and you’re leveraging it.’* **Case four**, Educator, Adult Hospice, White British

The type of community engagement activities participants described included visiting and working with local faith-based organisations, hosting religious and cultural events, attending external community events, using social media and targeted advertising within ethnically diverse areas. The importance of showing respect, reciprocity and relational humility during these interactions and rejecting a top-down or expert-dominated stance was highlighted:

> *‘And you always feel very humbled when you turn up and there’s dozens and dozens of people there to come and hear you talk about this because you might think that, oh, you know, is it what anyone wants to do on, like, a rainy Wednesday night going to talk? About death and dying, but obviously when you hear the stories and experiences people have, it kind of makes you realise how important it is, you know, to to a lot of people.’* **Case one,** Doctor, Hospital, White British

A key component of community engagement was services working alongside a *‘trusted voice’* (Spiritual care worker, Hospital, Asian/Asian British) in the local community. Faith and community leaders acted as trusted messengers within their communities, reduced fear and anxieties and increased acceptance by legitimising and sharing their knowledge of palliative care and the services available within the local area:

> *‘They can cascade that information down and then it’s coming from someone they trust or coming from someone within the community. And it’s much easier to get that access because it’s not an outsider coming and going coming in.’* **Case four**, Family support worker, Children’s Hospice, Black/Black British/Caribbean/African

Moving away from a westernised, one-size-fits-all explanation of palliative care towards a more person-led and culturally safe approach was recommended to reduce stigma and increase acceptance. Specialist palliative care professionals and referring clinicians recognising that different cultural relationships with death and dying exist and introducing the role of palliative care in ways that are accessible and meaningful to people’s communication needs was viewed as key:

> *‘If you’re believing that God is is here to or God will heal this child. So this child is not going to die. If there’s a miracle that could happen, that’s fine. So I’m just stating a practical example to you. So as a Hospice then then they can say to them well, that’s that’s OK. But in This is why we are here. So we are instrument you know, not to make things better but whilst we are waiting for that miracle to happen we are here to support you.’* **Case six,** Parent, Children’s Hospice, Black/Black British/Caribbean/African

Clearly communicating the additional support and opportunities available from accessing support could help patients and families understand palliative and hospice care as an enhancement to existing care, rather than a replacement or an indication of the end of life. Emphasising that hospice involvement could coexist with ongoing care, support life at home, and create meaningful experiences could support patients and families to see hospice care as relevant, supportive, and aligned with their values:

> *‘So what we wanted to do is have something at the Hospice with community leaders so they could then use their platform. To then you know, experience what the Hospice was all about. It’s not just that end of life experience at all. And there’s no shame in using in having help. It’s not. It’s not like we’re taking away the the care inside it. It’s just adding something.’* **Case four,** Hospice Director, Children’s hospice, White British.

The importance of services recognising that word of mouth and everyday encounters can influence trust and future engagement beyond the individual patient or family was highlighted. Experiences of palliative and hospice care can shape wider community perceptions with positive experiences helping to build trust and acceptance, but negative stories can also be quickly shared:

> *‘We hear good stories, as in good news and as you know, good news travels. But bad news travels quicker.’* **Case three,** Spiritual care worker, Hospital, Asian/Asian British

Engagement with palliative care services was seen to be influenced by generational differences in values, expectations, and experiences with younger generations perceived as more open to support. It was felt younger generations could play an important role as intergenerational bridges, supporting conversations within families and communities about hospice and palliative care based on shared experience and evolving perspectives:

> *‘So I think it is also that mindset kind of thing. And then as I think as generations get older, I think it will change. And I think that my generation, the next generation will be more knowledgeable in terms of what’s available.’* **Case five**, Patient, Adult Hospice, Asian/Asian British

### Workforce composition and diversity help to bridge the gap between ethnically diverse communities and services

A diverse workforce across all areas including leadership roles can help broker and bridge the gap between palliative care services and ethnically diverse communities. Patients and families seeing themselves in staff and volunteers who reflect the local community can create a sense of belonging, reduce apprehension and challenge assumptions about who services are for. A diverse workforce supports trust-building and legitimises care through familiarity and shared experience:

> *‘And it was a consultant or a doctor who was from the from a minority community so that was the bridge. So they said, look, you know, it’ll do you a lot of good, you know, all the support, there’s this, there’s this.’* **Case one,** Counsellor, Children’s Hospice, Asian/Asian British

In some areas there could be a lack of representation in leadership roles which participants felt needed to be addressed to facilitate equitable care:

> *‘Working in (place name) you just go into the (name of hospital) and you go into places and the staffing actually is diverse. But obviously I suppose the only thing I would say is that it’s diverse, But as how do I put this at certain levels? It’s good for people to see people like them at senior levels.’* **Case two,** Occupational Therapist, Hospital, White British

Staff from ethnically diverse communities supported and guided their colleagues on how to provide culturally safe care. Staff and volunteers from ethnically diverse communities facilitated and, in some cases, instigated the development of community and faith-based initiatives within their organisations and local communities to improve engagement and access:

> *‘And because we’ve got colleagues from the BAME communities who said actually we understand what this is about, we want to get it right. We don’t want it to be, you know, disrespectful. We want to do it in the right way. They supported us with the right engagement, the right foods, the right level of social media and the right interface. And it was a very, very successful evening.’* **Case four,** Manager, Children’s Hospice, White British

Volunteers embedded in local communities could encourage others to engage in palliative care services and shift understandings of hospice care that are shaped by limited or symbolic exposure, rather than direct knowledge of what is offered:

> *‘Because I did not know that all of these things that was taking place here. And there was no information out there that you could read about what was taking place unless you came in. That’s how you know, if you’re outside, you don’t know. You just saw a building just looking in. Oh, yeah, that, that over there. That’s where they die.’* **Case two**, Volunteer, Adult Hospice, Black/Black British/Caribbean/African

### How communication practices enable equitable care for people who have English as an additional language

The need for effective communication practices to support people who have English as an additional language was seen as imperative to enable equitable care and facilitate informed choice, especially at critical moments in the persons care pathway. Early identification of a person’s communication needs was seen as key to enabling safe and equitable care provision:

> *‘And I think that again has to start from the beginning and it’s just a simple tweak when the referral comes in or they have the first meeting to fill out the forms like we do with anything that we do. I think one of the boxes should be what is your primary language that you would prefer to communicate in or what language do you understand?’* **Case one**, Counsellor, Children’s Hospice, Asian/Asian British

Being able to access professional interpretation services was also seen as vital as it facilitated mutual understanding, with interpreters supporting staff to understand what mattered to patients and enabling patients and families to fully participate in conversations about their care and feel more at ease:

> *‘So the interpreters are very, very important. OK, because we can explain them clearly and they can explain to us, clearly.’ (via interpreter)* **Case one,** Parent (Bereaved), Children’s Hospice, Asian/Asian British

Multilingual staff also helped patients and families to navigate care systems, enabled their voices to be heard, and feel more at ease and engaged. Multilingual staff were supported by professional interpreters as appropriate for sensitive and complex medical conversations:

> *‘One of the children was so dissociated and as soon as I spoke in Arabic. You know, you know, they would light up again and they would be in the room again. And and I could see that physically, physically happening. You know, I could see it happening in front of me.’* **Case six,** Therapist, Children’s Hospice, Mixed/multiple ethnic group

The availability of high-quality translated materials in multiple languages and formats which can be accessed in public and clinical spaces was seen as imperative to support equitable access to information about services and reduce anxieties:

> *‘But others like they just speak their own language, Gujarati only or maybe Hindi only or whatever language they speak. If they don’t know, they won’t be able to know about (name of hospice) are doing this (name of hospice) are doing that. They won’t know if they get the information in English, they will just see. And they said I don’t understand. They’ll throw it away.’* **Case five,** Patient, Adult Hospice, Asian/Asian British

Accessibility was understood to involve more than translation alone, requiring materials to be co-reviewed with members of the intended communities to ensure meaning, imagery, language, and examples resonate with different cultural contexts:

> *‘How do Turkish people make sense of this as it is, or how people from I don’t know speaks Arabic makes sense of this what’s written, and this is something that was not really focused on before because once something is translated, oh, we’re being equitable, but actually not because. Just translating to another language does not mean it’s accessible to people.’* **Case two,** Nurse, Hospital, Other ethnic group

### Organisational commitment and partnership as drivers of sustainable change

Leaders within health care organisations prioritising and being committed to providing culturally safe care and working in partnership with other organisations was seen as a key driver of sustainable change. Organisations and funders needed to be accountable, demonstrate ownership and not just rely on isolated or short-term initiatives to address inequities in service. Leadership support at both board and clinical levels along with funding and resources to support the implementation of culturally adapted innovations into practice was imperative for sustainable change:

> *‘I think the trust is quite forward thinking with that there is, you know, that kind of appropriate care to our diverse communities is very much at the forefront of our Trust. So I think we’re lucky with that. It was, yeah, it was substantially funded’* **Case three,** Nurse, Hospital, White British

Cultural awareness and safety embedded in policies, staff development and cross sector partnership working were seen as key indicators of an organisations commitment to providing equitable care. Organisational policies and procedures that were responsive to cultural and faith practices could support trust between services and communities and make staff feel valued:

> *‘A child from a Muslim background pass away and everything was on track. You know they had everything. We’ve got all the resources because obviously they have to have a burial, which is in 24 hours. And they had everything. You know, the family was really grateful.’* **Case five,** Link worker, Children’s Hospice, Asian/Asian British

Organisations collecting and evaluating data on equitable access, delivery and clinical outcomes across ethnicities within their catchment areas was also imperative to identify gaps in service provision. Organisational priorities, strategy and delivery being informed by high quality data was seen as a key driver to enable sustainable transformative change:

> *‘What feels to me, key about [the service name] was understanding our local data and our local demographics. And where we were sort of… because we were an outlier nationally, we knew that there were more people being admitted locally in the last three months compared to other places. So understanding that, and trying to work out how to fill the gaps, rather than just duplicating existing services, creating something that actually filled those gaps.’* **Case three**, Doctor, Adult Hospice, White British

Organisations creating a learning culture through staff development and training opportunities was seen as vital to facilitate culturally safe and meaningful care. Staff described developing confidence through experiential learning and reflective and relational practices rather than through acquiring fixed cultural knowledge:

> *‘So having an open mind to learning and being genuinely curious and interested, if you’re interested in helping the person you’re working for, you need to be interested in what’s important to them.’* **Case six**, Counsellor, Children’s hospice, Asian or Asian British

These approaches could support care that prioritised respect, humility, and shared understanding rather than professional or institutional norms. Openly inviting patients and families to articulate their ethnicity, wishes, beliefs, and priorities was key to enabling culturally safe and meaningful care, but staff could lack confidence during these encounters and worry about causing offence:

> *‘I think people are worried about causing offence. By because of that, they don’t ask and I think it’s better to if you don’t know, ask. People aren’t offended if they say, oh, you’re this, you’ve got that religion down. What does that mean for you? Because just because someone’s Muslim, it’s there’s a whole range of what that means to them.’* **Case two,** Occupational Therapist, Hospital, White British

Embedding cultural awareness and safety in cross sector partnership working between hospital, community, hospice services and community organisations was also seen as a key driver of sustainable change.

## Discussion

### Summary of findings

The findings highlight the importance of organisations creating the conditions for culturally and spiritually safe care for those from ethnically diverse communities rather than developing formally funded (often short-term) initiatives (see supplementary file one for a definition of culturally safe care). Key were building engagement and trust between ethnically diverse communities and the providers that serve them as a mechanism to promote access; workforce composition and diversity to enable building bridges between ethnically diverse communities and services; how communication practices can enable equitable care for people who have English as an additional language and organisational commitment and partnership as drivers of sustainable change. The final theoretical propositions are presented in table 2.

**Table 2.** Final theoretical propositions.

| Proposition | Description | Assumptions |
| --- | --- | --- |
| Proposition 1: Creating the conditions for culturally safe care | Culturally safe palliative care is more likely to be achieved when organisations intentionally create structural, environmental, and relational conditions that enable flexibility, dignity, and recognition of diverse beliefs and practices. | Cultural safety does not arise solely from individual clinician competence, but from organisational arrangements. These include the policies, environments, leadership and norms that guide organisations. |
| Proposition 2: Engagement and trust building as mechanisms that promote access | Sustained engagement and trust-building between ethnically diverse communities and service providers functions as a generative mechanism that increases awareness, reduces perceived risk, and promotes earlier and more equitable access to palliative care. | Trust is an outcome of repeated, credible, and reciprocal interactions. Where providers invest in long-term relationships understanding about palliative care is enhanced, referral pathways are improved, and communities have the information needed to facilitate timely access to services. |
| Proposition 3: Workforce composition and diversity as a means of bridging the gap between services and families. | A diverse workforce, particularly where staff share linguistic, cultural, or experiential backgrounds with local communities, enables brokerage between ethnically diverse populations and services. This facilitates access, navigation, and mutual understanding. | Workforce diversity operates not only symbolically but functionally. Staff members may act as cultural brokers. This may include translating expectations, supporting shared understanding, and legitimising services within communities. The visibility of staff from local ethnically diverse communities |
|  |  | in senior positions can increase trust and acceptance. |
| Proposition 4:<br>Communication practices as enablers of equitable care for people with English as an additional language | High-quality, structured communication practices enable equitable participation in decision-making and improve the quality and safety of palliative care for people who have English as an additional language | Where services ensure reliable interpretation, translated materials, and staff competence in working with interpreters, patients and families are better able to express preferences and understand options. |
| Proposition 5:<br>Organisational commitment and partnership as drivers of sustainable change | Sustainable improvements in equitable palliative care are most likely when organisational leadership demonstrates explicit commitment to equity and forms structured partnerships across health, social care, and community sectors. | Isolated initiatives are insufficient for durable change. Sustained impact depends on leadership endorsement, allocation of resources, integration into governance and performance frameworks, and formalised partnerships with community and voluntary organisations. |

### Relationship with wider theory and empirical work

We have used an adaptation of Bronfenbrenner’s Ecological Systems theory (Bronfenbrenner, 1994) and its five interrelated systems to highlight study implications, with particular reference to health policy. The interrelationship of this theory and our core findings is illustrated in Figure 1.

**Figure 1.**
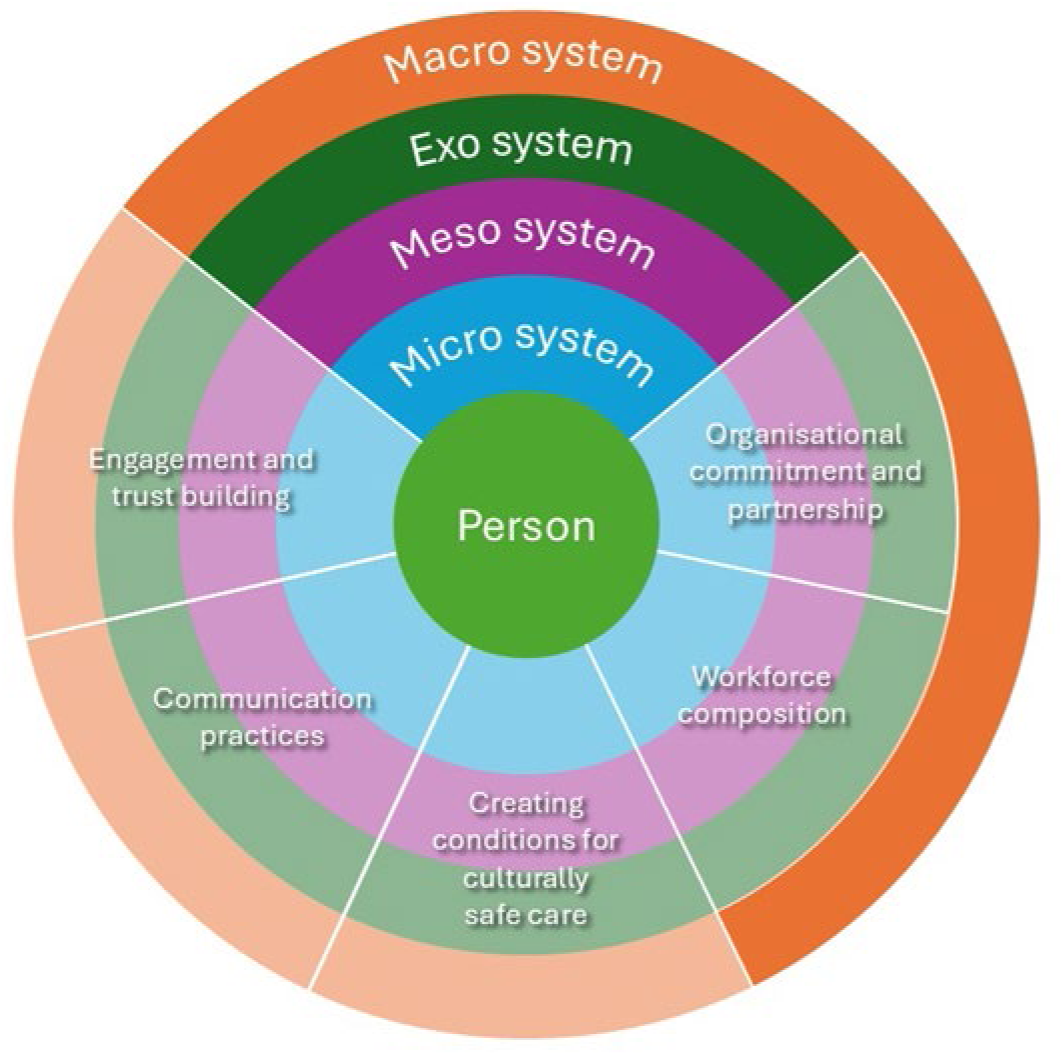
A representation of the interplay between the ecological systems theory and the thematic findings of this study.

#### The micro and mesosystems – the individual, their social networks, and the interactions between them

At the microsystem level, the experience of palliative care is shaped by people’s immediate relationships and everyday interactions. The quality of these encounters appears central to gaining and maintaining trust, enabling partnership and facilitating shared decision-making. Language and culturally mediated understandings of illness, dying and prognosis influence how information is received and acted upon (Hira et al., 2025). Communication challenges, including limited access to professional interpretation, can compound uncertainty and distress and continue to shape the experiences of many patients and their families (Clarke et al., 2023). Policy actions at provider levels are needed at this microsystem level with a focus on addressing communication challenges, improving the quality of interactions, and providing person-led care.

The mesosystem concerns the interactions and coordination between these microsystem actors. Fragmented partnerships and culturally unsafe practice can restrict access and undermine continuity. Conversely, collaborative working grounded in cultural humility and safety strengthens links between services and communities, enabling care that is responsive rather than reactive (Curtis et al., 2019). From this ecological perspective, improving access to palliative care is not solely about individual practitioner competence, but about strengthening the relational bridges between families, professionals and community networks that collectively shape the end-of-life experience.

#### The exosystem – systems, services and policies that indirectly influence and affect the care a person receives

Clinical and service leaders need to proactively embed cultural safety into their physical spaces and policies to build trust and increase acceptance (Fisher et al., 2021; Walton, 2025). Providing and implementing faith specific resources, including access to faith leaders, and policies that allow staff to support cultural and religious practices is imperative to address inflexible care routines, enable meaningful care, and signal inclusivity (Klitzman et al., 2023).

Specific health policy needs were identified in areas such as interpretation, learning environments and the use of service-related data. First, organisations must be able to offer high-quality professional interpretation services and co-designed translated materials in multiple languages and formats to ensure equitable and safe care for people who have English as an additional language (Bigger et al., 2025; Lokugamage et al., 2023). The needs of interpreters operating in such a sensitive and challenging area as palliative care must not be overlooked (Hancox et al., 2023; Latif et al., 2023), and care professionals trained in how to work effectively alongside interpreters (Green et al., 2018; Latif et al., 2022). Second, facilitating cultural safety involves leaders creating a learning environment where cultural humility is central to the process and where power imbalances and historical negative experiences are recognised (Hussain et al., 2025; Lokugamage et al., 2023). Mandatory training should focus on promoting curiosity rather than tick box learning (Bansal, 2016). Third, services need to collect standardised high quality data that measures equitable access and delivery, and whether there are equal outcomes among the population they serve (Chidiac et al., 2025).

### The macrosystem: societal attitudes and culture

This system relates to wider societal attitudes, including systemic racism, culture and customs such as those related to death and dying and how palliative care services are structured and funded. Services need to move beyond their institutional boundaries and proactively engage with ethnically diverse communities using an asset-based approach to reduce fear and improve acceptance of palliative care (NHS England, 2022b). This involves identifying and bringing together existing community values, resources and expertise rather than identifying and ‘fixing’ deficits from the top down (Matthiesen et al., 2014). Services should consider providing care within culturally meaningful spaces and partnership activities should focus on building links with community and faith organisations and leaders to build trust and reduce fear through mutual learning and co-design of services (Ip et al., 2026). Multi agency buy in and extra resources are required to implement and sustain more formal engagement and partnership initiatives (Roleston et al., 2023).

The issue of workforce diversity as a mechanism to improve access is a complex issue. Diversity in the workforce does not automatically lead to high levels of cultural appreciation with an organisation. Clinical and service leaders need to engage in meaningful recruitment that reflects the local population rather than tick box diversity recruitment (Khiroya & Willis, 2022). Having individuals in senior positions that reflect the local community they serve to facilitate and sustain transformational change to ensure equitable and culturally safe care is important. Engaging volunteers of all ages, including young people, from diverse backgrounds can foster community wide partnership and facilitate shared learning (Shinan-Altman, 2026).

### The chronosystem: change and transition over time

In palliative care the temporal dimension is crucial to understand how care is accessed and experienced across the life course. Temporality in palliative care provision also reflects an urgency given the potential anticipated deterioration and death of the palliative care population making compassionate, coordinated, and person-led care essential from the outset. Specific examples of temporality include the provision of palliative care through a range of developmental stages (Iluno et al., 2024; Persaud et al., 2026), and how it affects care as people age and are multiply bereaved. Individual life transitions such as migration can also influence when and how people encounter palliative care services. (De Souza et al., 2024).

Shifts in public discourse around race, evolving models of culturally competent practice, and changing migration patterns all influence service design and accessibility. Younger generations may hold different expectations of autonomy and advance care planning, creating intergenerational tensions within families (De Souza et al., 2024). Recognising these temporal dynamics enables palliative care to be responsive not only to cultural identity, but to how identity, trust, and need evolve over time.

## Strengths and limitations of the research

The findings reflect participants perspectives across a range of areas and settings within the context of adult and paediatric palliative care and focus on solutions relevant to health policy, rather than the barriers that have been previously studied. Some of the findings are unique to palliative care, but others may be transferable to other areas. The immediate research team worked alongside PPIE and academic colleagues from ethnically diverse communities to ensure the research was conducted with cultural sensitivity (Elliott-Button et al., 2026).

We were unable to recruit a case that included a large Chinese or East Asian population and the majority of patient and family carer participants identified as Asian or Asian British so the views of other communities may not be fully represented. The experiences of those who live in areas with smaller ethnically diverse communities, or in rural or other isolated localities may not also have been represented. We collected limited documentary data due to the small number of formal initiatives within the cases.

## Conclusion

Equitable access to high-quality palliative care is not a marginal policy issue; it is indicative of the values that underpin our health and care systems. The evidence is clear and well-known: people from ethnically diverse communities continue to experience later referrals to and use of palliative care services, poorer communication, and have unmet needs towards the end of life. It is likely that many of these issues related to access and experience are outcomes of systems that were not designed with all communities in mind.

Examples of community engagement, cultural knowledge, and voluntary and faith-based organisations already engaged in supporting people at the end of life were identified. The challenge is not necessarily the absence of community capability, but how health and care systems recognise, partner with, and build upon existing strengths. Changes require intent, accountability and leadership. Critically, improving care is likely to depend on reallocating attention and responsibility. Clear accountability mechanisms, metrics linked to equity, and expectations that services demonstrate responsive and appropriate palliative care could shift practice.

The economic argument for addressing inequities is also strong. Late crisis admissions, high use of emergency care, poorly coordinated care and avoidable hospital deaths are costly for the system and often distressing for families. Earlier, equitable access to palliative care is likely to improve symptom control, support carers and reduce unplanned acute care use. There is also a clear moral imperative to act. Palliative care is concerned with dignity, comfort and respect at the most vulnerable point in a person’s life. A health and care system committed to fairness cannot look away from disparities that shape how people die and how families remember those final days.

## Acknowledgements

Patient and Public Involvement Group members Rita Antonova, Zahida Aslam, Saima Gull, Patricia Jamal, Emily Lam, Jenny Richards, Ms Farheen Yameen

This research is funded through the NIHR Policy Research Unit in Palliative and end of life care, reference NIHR206122. The views expressed are those of the author(s) and not necessarily those of the NIHR or the Department of Health and Social Care.

Grant funding for research but no other competing interest: All authors have completed the ICMJE uniform disclosure form at www.icmje.org/coi_disclosure.pdf and declare: all authors had financial support from the NIHR Policy Research Unit in Palliative and End of Life Care (grant number NIHR206122) for the submitted work and Professor Fraser also had financial support from the NIHR Academy (grant number NIHR304252) for a NIHR Research Professor Award; no financial relationships with any organisations that might have an interest in the submitted work in the previous three years; no other relationships or activities that could appear to have influenced the submitted work.

For the purpose of Open Access, the author has applied a Creative Commons Attribution (CC BY) licence to any Author Accepted Manuscript version arising.

## Data statement

Due to the sensitive nature of this study, participants were assured that their non-anonymised data would remain confidential and would not be shared.

## CRediT statement

Conceptualization SB, CE, LF, SM, NP, CW

Data curation LD, SG, GC, NC, CW

Formal analysis LD, SG, GC, NC, CW

Funding acquisition SB, CE, LF, SM, NP, CW

Investigation LD, SG, GC, NC

Methodology SB, CE, LF, SM, NP, CW

Project administration LD, SG, GC, NC, CW

Resources N/A

Software N/A

Supervision CW

Validation N/A

Visualization LD, SG and CW

Writing – original draft- LD, SG and CW

Writing – review and editing LD, SG, GC, SB, CE, LF, SM, NP, CW

