## Supplementary file 1 for "Improving palliative care for babies, children, young people and adults from ethnically diverse communities: a qualitative multiple case study"

**Additional stakeholder, patient and family carer participant demographic information**

| | **Case One (n=9)** | **Case Two (n= 8)** | **Case Three (n=20)** | **Case Four (n=10)** | **Case Five (n=18)** | **Case Six (n=6)** |
|---|---|---|---|---|---|---|
| Health and Social Care professionals including volunteers | | | | | | |
| Gender | F=7 | F=6<br>M= 2 | F=9<br>M= 2 | F=9<br>M= 1 | F=8 | F=3<br>M= 1 |
| Country of birth | UK=7 | UK=3<br>Non-UK=5 | UK=9<br>Non-UK=2 | UK=10<br>Non-UK=0 | UK=6<br>Non-UK=2 | UK=2<br>Non-UK=2 |
| Ethnic Group | White British= 4<br>Asian/Asian British =3 | White British= 4<br>White other=2<br>Black, Black British, Caribbean or African=1<br>Other ethnic group=1 | White British= 5<br>Asian/Asian British =5<br>Other ethnic group=1 | White British= 6<br>Asian/Asian British =3<br>Black, Black British, Caribbean or African=1 | White British= 5<br>Asian/Asian British =2<br>Black, Black British, Caribbean or African=1 | White British=1<br>Asian/Asian British =2<br>Mixed/multiple ethnic group= 1 |
| Main Language | English=7 | English=8 | English=9<br>Arabic=1<br>English & Urdu=1 | English=8<br>Gujarati=1<br>Gujarati and English=1 | English=5<br>Gujarati and English=2<br>Shona=1 | English=2<br>English/Cantonese =1<br>French=1 |
| Religion | No religion= 3<br>Muslim=3<br>Christian=1 | No religion= 6<br>Christian=2 | No religion= 3<br>Muslim=4<br>Christian=2<br>Sikh=1<br>Hindu=1 | No religion= 2<br>Muslim=3<br>Christian=5 | No religion= 3<br>Christian=3<br>Hindu=2 | No religion= 1<br>Muslim=1<br>Christian=1<br>Hindu=1 |

| | | | | | | |
|---|---|---|---|---|---|---|
| Highest level of education | Bachelors/Masters= 4<br>High/secondary school/diploma= 3 | Doctoral=4<br>Bachelors/Masters =3<br>High/secondary school/diploma= 1 | Bachelors/Masters =9<br>High/secondary school/diploma= 2 | Bachelors/Masters =9<br>High/secondary school/diploma= 1 | Bachelors/Masters =6<br>High/secondary school/diploma= 2 | Bachelors/Masters =3<br>High/secondary school/diploma= 1 |
| Primary professional role | Doctor=1<br>Nurse=3<br>Counsellor= 1<br>Family support worker=1<br>Volunteer=1 | Doctor=1<br>Nurse=3<br>Occupational therapist = 1<br>Psychologist=1<br>Spiritual care worker=1<br>Volunteer=1 | Doctor=3<br>Nurse=4<br>Spiritual care worker=1<br>Manager/admin=2<br>Link worker=1 | Doctor=1<br>Nurse=2<br>Family support worker=1<br>Spiritual care worker=1<br>Manager/admin=2<br>Educator=1<br>Volunteer=2 | Nurse=3<br>Family support worker=1<br>Volunteer=1<br>Manager/admin=1<br>Link worker=1<br>Key worker=1 | Nurse=1<br>Counsellor=1<br>Therapist=2 |
| Primary work Environment | Hospital= 4<br>Hospice= 3 | Hospital= 5<br>Hospice= 2<br>Community=1 | Hospital= 3<br>Hospice= 5<br>Community=3 | Hospital=3<br>Hospice= 6<br>Community=1 | Hospice= 6<br>Community=2 | Hospice=4 |
| Specialist/ generalist palliative care or other primary role | Specialist=5<br>Generalist=2 | Specialist=5<br>Generalist=3 | Specialist=10<br>Other primary role=1 | Specialist=5<br>Other primary role=5 | Specialist=5<br>Generalist=1<br>Other primary role=2 | Specialist=1<br>Generalist=2<br>Other primary role=1 |
| Adult or Children's services | Adult=4<br>Children=3 | Adult=8 | Adult=11 | Adult=6<br>Children=3 | Adult=3<br>Children=5 | Children=4 |

| Patients and family carer participants | | | | | | |
|---|---|---|---|---|---|---|
| Gender | M=1<br>F=1 | | M= 4<br>F=5 | | M= 4<br>F=6 | M= 1<br>F=1 |
| Age Group | 31-40=1<br>41-50=1 | | 31-40=3<br>41-50=4<br>52-60=1<br>71-80=1 | | 31-40=2<br>51-60=4<br>61=70=4 | 18-30= 1<br>41-50=1 |
| Country of birth | UK= 1<br>Non-UK =1 | | UK=5<br>Non-UK=4 | | UK=3<br>Non-UK=7 | Non-UK=2 |
| Ethnic Group | Asian/Asian British =2 | | Asian/Asian British=6<br>Other ethnic group=1<br>White other=1<br>White British=1 | | Asian/Asian British=8<br>Mixed/multiple ethnic group= 1<br>Black, Black British, Caribbean or African=1 | Black, Black British, Caribbean or African=1<br>White other=1 |
| Religion | Muslim=2 | | Muslim=5<br>Hindu=3<br>Christian=1 | | Hindu=6<br>Muslim=2<br>Christian=2 | Jewish=1<br>Christian=1 |
| Main Language | English=1<br>Urdu=1 | | English=7<br>Arabic=1<br>Portuguese=1 | | English=5<br>Hindi/English=1<br>Mandarin=1<br>Gujarati=1<br>Hindi=1<br>Urdu=1 | Ga=1<br>Romanian=1 |
| Highest level of education | Primary=1<br>Bachelors/Masters= 1 | | Bachelors/Masters =7<br>High/secondary school/diploma=1<br>No education=1 | | Doctoral=1<br>Bachelors/Masters =5<br>High/secondary school/diploma=4 | Bachelors/Masters =2 |

| | | | | | | |
|---|---|---|---|---|---|---|
| Diagnosis | Genetic condition=2 | | Cancer= 3<br>Non-cancer=4<br>Both=2 | | Cancer=4<br>Non-cancer=5<br>Genetic condition=1 | Congenital condition=1<br>Epilepsy=1 |

**Initial theoretical propositions (Context-mechanism-outcome configurations)**

| | |
|---|---|
| **CMOc 1: The impact of structural racism on trust and engagement in services** | Historical and institutional racism, including direct and indirect experiences of racism, can result in people from ethnically diverse communities receiving inadequate care that fails to meet their cultural needs (C). This can lead to a loss of trust (M) in services and fear (M) of poor treatment, resulting in non-engagement with palliative care and a preference for seeking support within their own communities (O). Such effects may be heightened at the end of life, a time of increased vulnerability (C). |
| **CMOc 2: The cultural significance and importance of family being present and actively involved in end-of-life decision making** | The cultural significance of family amongst many people from ethnically diverse communities means that families often seek active involvement in end-of-life decision-making (C). This can enhance the sense of autonomy for both the patient and family, particularly when the patient trusts their family to honour their wishes (M). However, conflicts within families (M) may arise, leading to disagreement between family members about whether, for example, to withhold information from the patient about their prognosis. This can result in the patient's voice being overlooked and create feelings of ethical unease among healthcare staff (O). |
| **CMOc 3: Promoting cultural sensitivity, inclusivity and awareness of palliative care.** | Palliative care settings often lack cultural and religious understanding, such as having inpatient policies that fail to prioritise the need for family being present at the end of life, as well as services failing to reach and engage with people from ethnically diverse communities (C). This can lead to people from ethnically diverse communities feeling excluded (M) and creates a lack of awareness (M) about |

| | |
|---|---|
| | available service options, resulting in non-engagement (O) and a preference among some to die at home rather than in healthcare settings. |
| **CMOc 4: The importance of services delivering culturally safe and sensitive care** | (C) Healthcare staff can deliver culturally safe, person-centred care when their practice is informed by an understanding (M) of cultures other than their own. This fosters feelings of safety (M) for people from ethnically diverse communities and their families, empowerment (M), and trust (M), in-turn improving staff-patient relationships (O) and leading to higher satisfaction for people from ethnically diverse communities and their families (O). |
| **CMOc 5: Challenges of understanding concepts specific to palliative care** | The absence of suitable translation, interpretation, or understanding of the concept of palliative care and/or palliative care terminology (e.g. DNACPR) makes it difficult for people from ethnically diverse communities with limited English proficiency to understand and consent to care (C). Thus, patients from ethnically diverse communities and their families may feel the services are ‘not for them’ or they may lack knowledge of what available services include (M). This may lead them to self-manage or seek guidance from religious leaders instead of accessing services (O), resulting in delayed engagement with palliative care, often only as emergency hospital admissions at advanced stages of illness (O). |

**Definition of Cultural Safety** (taken from Curtis E, Loring B, Jones R, et al. Refining the definitions of cultural safety, cultural competency and Indigenous health: lessons from Aotearoa New Zealand. *Int J Equity Health* 2025; 24: 130.)

The term cultural safety refers to the process where practitioners and service providers hold themselves accountable by actively engaging in ongoing self-reflection and self-awareness to assess the potential impact of their own culture on clinical interactions and care delivery. Biases, prejudices, attitudes, stereotypes and assumptions as well as the structures and characteristics that may affect care quality need to be acknowledged, addressed and monitored. It is important to develop culturally safe organisations and not just focus on individual responsibility. Importantly patients, family carers and their social networks define whether care provision is culturally safe. Power needs to be transferred to patients and communities to comment on the safety of their care experiences from a cultural perspective.
